# Factors Associated with the Deterioration of Post-COVID-19 Condition Symptoms Following a Dose of SARS-CoV-2 Vaccine

**DOI:** 10.64898/2026.01.06.26343459

**Authors:** Malika Seydou Beidari, Martyne Audet, Stéphane Turcotte, Raoul Daoust, Corinne M. Hohl, Patrick M. Archambault, Canadian Emergency Department Research Network, Network of Canadian Emergency Researchers, Canadian Critical Care Trials Group investigators Canadian COVID-19 Emergency Department Rapid Response Network

## Abstract

**Objectives:** To investigate the factors associated with the deterioration of post-COVID-19 condition (PCC) symptoms in patients who received a dose of SARS-CoV-2 vaccine ≥ 90 days after their infection.

**Method:** Multicenter cohort study conducted in 33 emergency departments across Canada, including 476 patients who developed PCC according to the World Health Organization definition. Data were collected via telephone questionnaires. Statistical analyses, including logistic regression models, were performed to identify factors associated with symptom deterioration after vaccination.

**Results:** Among participants, 28.8% reported a deterioration of their PCC symptoms after vaccination. Two factors were significantly associated with this deterioration: receiving the Moderna (mRNA-1273) vaccine (aOR = 1.80; 95% CI: 1.14–2.8) and a persistent cough three months after the initial infection (aOR = 1.81; 95% CI: 1.03–3.15). No association was found between symptom deterioration and sociodemographic characteristics such as age or sex.

**Conclusion:** Post-infection vaccination may be associated with increased risk of PCC symptoms deterioration in some patients, particularly those vaccinated with Moderna (mRNA-1273) or presenting with a persistent cough. While the benefits of vaccination remain substantial, these findings call for further investigation into the underlying immunological mechanisms and possible adjustments to vaccination strategies for patients with PCC.

## Background

From the beginning of the COVID-19 pandemic, many patients reported experiencing persistent symptoms long after the acute phase of the infection, a condition now known as Long COVID or post-COVID-19 condition (PCC). In October 2021, the World Health Organization (WHO) defined PCC as the presence of at least one new symptom occurring within 3 months of a proven or suspected acute infection with severe acute respiratory syndrome coronavirus 2 (SARS-CoV-2) still present at 3 months and lasting for more than 2 months.^1^ Characterized by a myriad of often nonspecific symptoms that can last for months or even years after the initial infection, PCC can significantly affect patients’ quality of life.^2–4^ In the absence of distinctive biomarkers ^5^, the diagnosis of PCC remains difficult to establish and poses challenges for both the affected population and healthcare systems.^6,7^ Despite active research to elucidate the pathophysiology of PCC, the inflammatory pathway, associated with tissue damage, organ damage, or an autoimmune response, remains one of the most studied potential pathophysiological pathway.

The introduction of SARS-CoV-2 vaccination in late 2020 marked a turning point in the pandemic. Subsequently, hypotheses have emerged suggesting that vaccines may have an impact on the progression of PCC symptoms.^8^ While vaccines have demonstrated their effectiveness in reducing the incidence of the disease ^9,10^, virus transmission ^11^, hospitalizations and deaths due to COVID-19 ^12^, and the development of PCC ^13,14^, few studies have been conducted on the impact of vaccination administered after a SARS-CoV-2 infection on established PCC symptoms.^15–17^ The existing literature on this issue is inconsistent. Recent studies suggest variable and contradictory impacts, such as improvement, deterioration, or no change, on persistent COVID-19 symptoms.^18–22^ Additional research is needed to understand whether stimulation of the immune system through vaccination has an impact on PCC symptoms in patients who met the definition of PCC 3 months after infection.

In this study, we investigated the association between SARS-CoV-2 vaccination and the deterioration of PCC symptoms among patients who met the WHO PCC criteria and were vaccinated at least 90 days after their initial infection.

## Methods

### Study design and setting

The Canadian Emergency Department Rapid Response Network (CCEDRRN), a Canada-wide collaboration including public health partners, aimed at standardizing the collection of data on patients tested for COVID-19 in participating emergency departments. This initiative included more than 50 emergency departments across eight provinces in Canada.^23^

This study was conducted in 33 of the 50 CCEDRRN sites across five Canadian provinces (Quebec, British Columbia, Nova Scotia, Saskatchewan, and Ontario). The study was approved by the research ethics committees of all participating institutions, which granted permission to contact patients to obtain verbal consent to participate in this research. This study was reported in accordance with the STROBE guidelines for observational studies (Appendix 1).^24^

### Participants

We recruited consecutive patients aged 18 years or older who visited a participating emergency department between October 18, 2020, and February 28, 2022, and who tested positive for SARS-CoV-2 (Appendix 2). All eligible patients were contacted by telephone after discharge from the emergency department or hospitalization and invited to participate in the study. Consenting participants completed at least one questionnaire on PCC symptoms, the *Post COVID-19 Condition Assessment Questionnaire (*PCCAQ) (Appendix 3), between November 19, 2021, and July 31, 2022, at 6 and 12 months after their emergency department visit. Participants had to have reported at least one symptom consistent with the WHO criteria for PCC and received at least one dose of vaccine ≥ 90 days after their visit to the emergency department. This criterion was applied to ensure the inclusion of only patients who had met the WHO criteria for PCC before receiving the reference dose of a COVID-19 vaccine. At the end of the questionnaire, we asked patients who had received a COVID-19 vaccine after an initial infection if they had noticed a change in their new persistent symptoms. We excluded participants who responded that they had not been vaccinated 90 days after the infection, who chose not to answer the question, or who could not answer the question because they were unsure or undecided about changes in their symptoms.

### Definitions

We defined SARS-CoV-2-positive patients as those with a positive nucleic acid amplification test or rapid antigen test at the emergency department index visit or from a sample collected in the community within 14 days prior to the index visit who remained symptomatic of acute COVID-19 at the time of their index visit.^23^ Hospitalized patients were included in this definition when their sample was collected within 14 days of arrival to the emergency department, reflecting the maximum incubation period for SARS-CoV-2 infection.

Based on the WHO clinical case definition, we defined meeting clinical PCC criteria as reporting: (1) at least one new PCC-consistent symptom arising in the 3 months after the emergency department visit that were present at the 3-month mark, and (2) lasted ≥2 months.^25^ PCC symptoms included dyspnea, pain, cough, loss of smell and taste, sleep disturbances, dizziness or vertigo, difficulty concentrating, memory problems, and post-exertional malaise. Other new symptoms since the emergency department visit were also considered based on the WHO case report form.^26^

The reference vaccination dose was defined as the first dose administered ≥ 90 days after the index visit to the emergency department, regardless of the total number of doses previously received. Vaccination status data were self-reported by participants. The validity of self-reported vaccination status and date of administration by patients in our database has already been presented in a previous study.^27^

### Data collection

Trained research assistants: (1) abstracted data on SARS-CoV-2 tested patients including their baseline comorbidities by chart review ^23^, (2) attempted to contact patients up to five times to obtain consent for phone follow-up 6 months and 12 months after the emergency department visit, (3) collected sociocultural and demographic variables including age, sex, self-reported gender, self-reported race, and self-reported SARS-CoV-2 vaccination status (number of doses received ≥ 7 days before ED index visit, dates of vaccination, and vaccine types), (4) documented any self-reported new or repeat SARS-CoV-2 infections, and (5) documented ongoing or resolved symptoms consistent with PCC using the PCCAQ (Appendix 3). Research assistants were instructed not to use the terms Long COVID or post-COVID-19 condition when questioning patients about any ongoing symptoms.

### Primary outcome and associated risk factors

The primary outcome was the deterioration of symptoms after vaccination, as self-reported by participants. To address limitations in sample size, we grouped participants who reported improvement with those who observed no change (status quo) in a no deterioration group and those who deteriorated in the deterioration group. This approach is commonly used in studies where the primary objective is to measure the change in disease status, or to prevent the deterioration of a chronic disease (e.g., cancer treatment ^28^, multiple sclerosis ^29^), or when the objective is to measure change in status in the course of cancer treatment.^30^ Since overall stability and improvement in PCC symptoms constitute an acceptable category of disease outcomes for patients, this approach was deemed appropriate.

### Statistical analyses

The selection of candidate predictor variables was based on a review of the literature, clinical acumen, the experiences of patient partners, and the results of previous studies.^15,31–34^ Thirteen variables were selected, including age, sex at birth, medical history, self-reported race, acute symptoms of COVID-19, self-reported symptoms of PCC, and type of vaccine received. The sociodemographic characteristics of the participants were compared using Student’s t-test and the chi-square test. Student’s t-tests were used to compare continuous variables, checking the assumptions of normality and homogeneity of variances. The chi-square test was used to assess associations between categorical variables including the primary outcome measure. We used multivariate logistic regression to identify independent factors associated with deterioration of PCC symptoms. We included variables with p-values ≤ 0.2 in bivariate analyses into the logistic regression model to retain as many potentially predictive variables as possible in the model. This approach was intended to minimize the risk of prematurely excluding variables that could play a role in the final model.

Subsequently, some variables were removed due to their small sample sizes. By excluding these variables, we sought to optimize the quality of the model fit while maintaining a balance between its complexity and robustness. The Hosmer-Lemeshow test was used to assess the overall goodness of fit of our model. We calculated odds ratios (ORs) with their 95% confidence intervals (95% CI) for each risk factor associated with the deterioration of self-reported symptoms by patients. We used SAS (version 9.4) to calculate descriptive statistics and logistic regression models.

### Patient partner involvement

For this project, the participation of CCEDRRN patient partners was limited to the development of the PCCAQ questionnaire. This questionnaire was developed based on the WHO PCC case definition and the WHO case report form, in collaboration with patient partners, PCC experts, emergency physicians, rehabilitation specialists, and public health decision-makers. We piloted the PCCAQ in English and French with patient partners and the first 100 participants.^35^ Patient partners did not participate in the writing of the current manuscript and were not involved in the interpretation of the results.

## Results

Of the 3,934 individuals assessed for eligibility, 1,532 had developed PCC. Of these, 533 received a vaccine dose ≥90 days after the index emergency department visit. Excluding participants who did not answer the PCCAQ question about change in their symptoms after receiving a vaccine dose (Question 16-A, Appendix 3), the final sample consisted of 476 participants (Figure 1).

**Figure 1:**
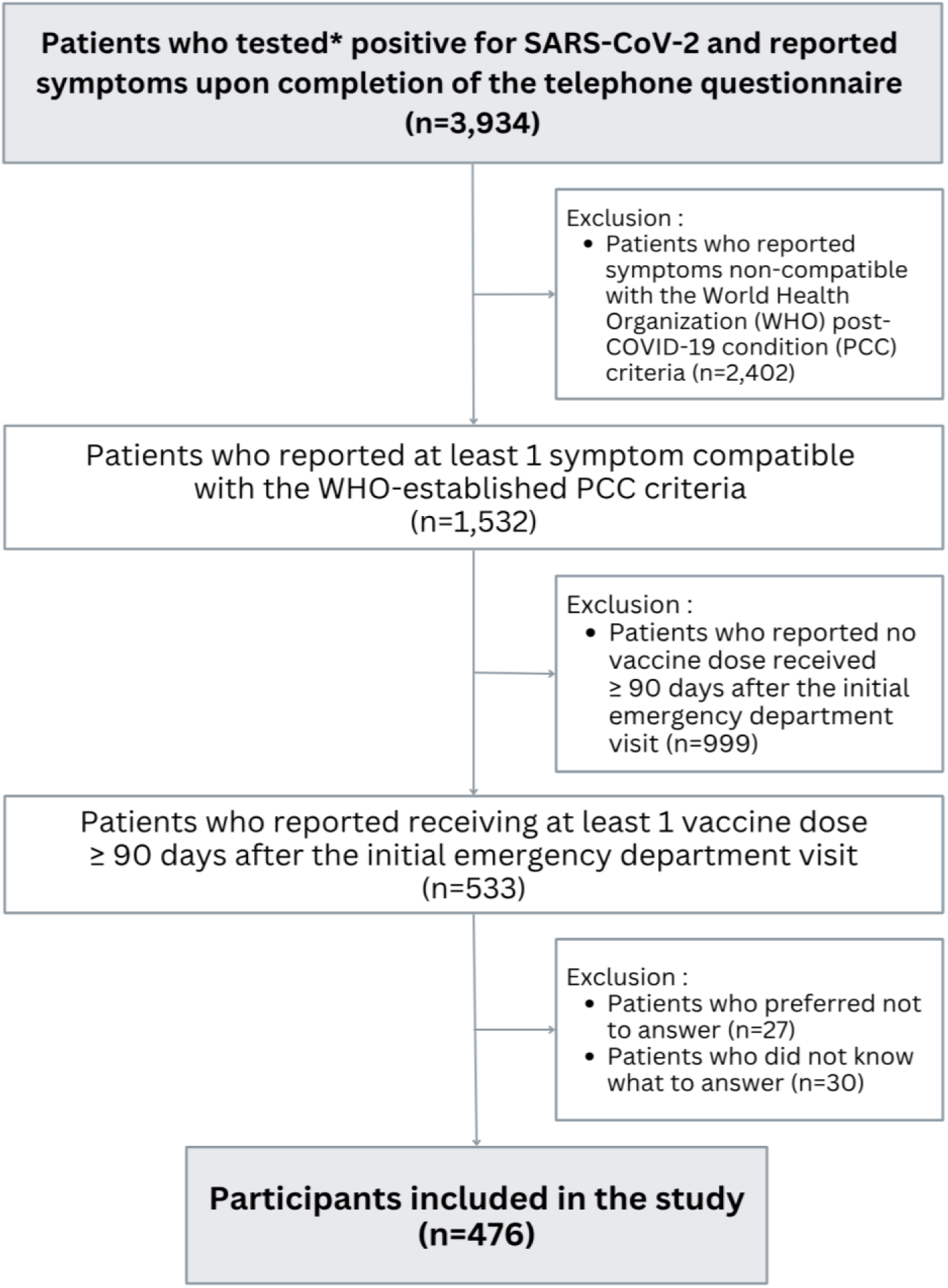
Flow chart of participants selected for the study. SARS-CoV-2: severe acute respiratory syndrome coronavirus 2, WHO: World Health Organization. *Participants who had a confirmed infection by nucleic acid amplification test (NAAT) or rapid antigen detection test (RAT) at the emergency department index visit.

Among participants, 28.8% (137/476, 95% CI: 24.7-32.9%) reported that their symptoms deteriorated after vaccination. Participants who reported improvement (10/476) or no change (329/476) accounted for 71.2% (339/476, 95% CI: 67.1-75.3%) of the cohort. Patients who deteriorated had similar baseline characteristics and acute symptoms of COVID-19 at their index emergency department visit compared to patients who did not deteriorate (Table 1). However, a higher proportion of participants who had developed a persistent cough (21.9%, 95% CI: 14.9-28.8%) and concentration difficulties (43.8%, 95% CI: 35.4-52.1%) 3 months after their index visit reported symptom deterioration after receiving a vaccine dose ≥90 days after their visit (Figure 2).

**Figure 2.**
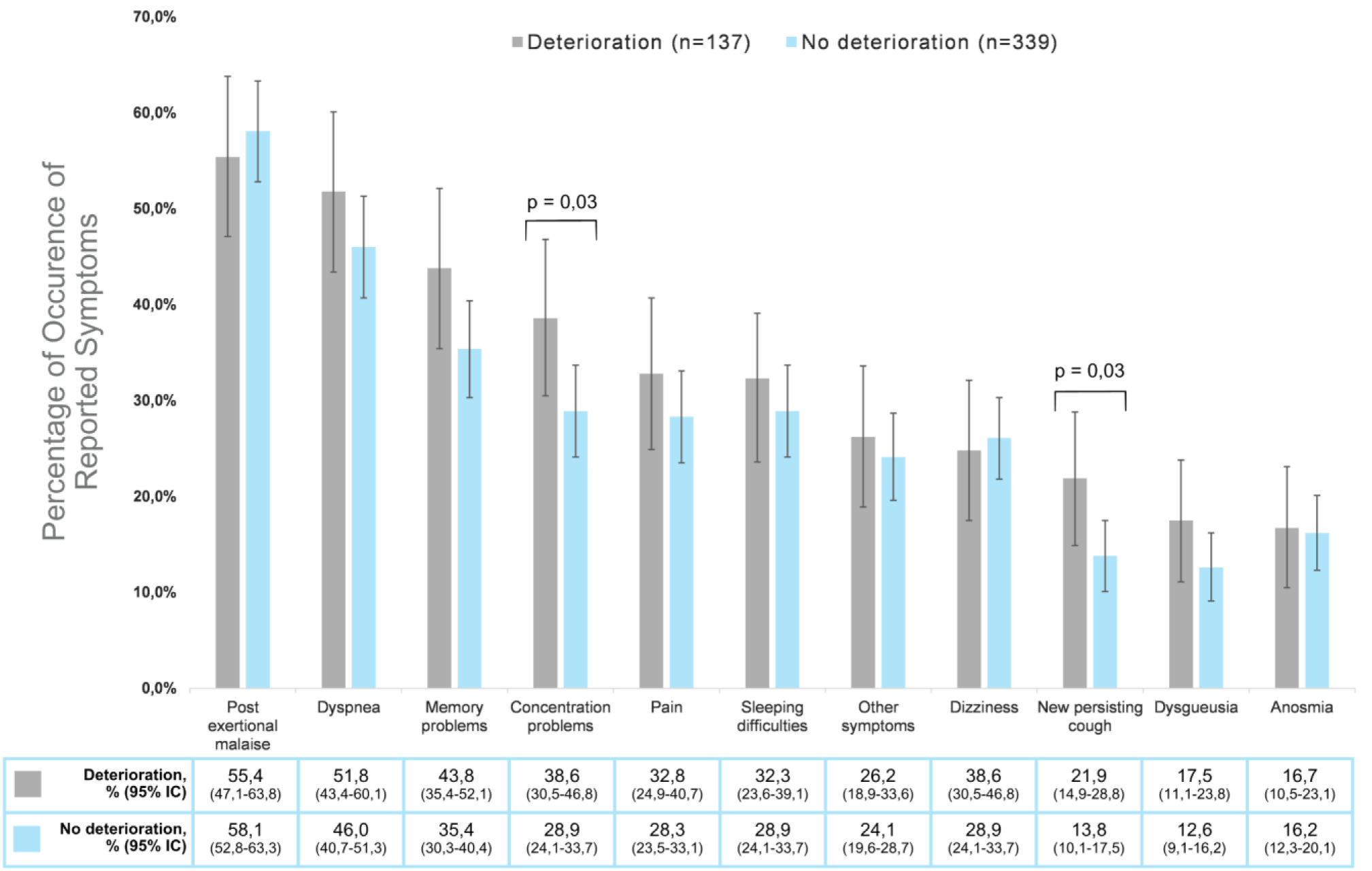
Symptoms of post-COVID-19 condition (PCC) reported by participants, stratified by symptom progression (deterioration versus no deterioration) after vaccination ≥ 90 days post-infection. (N=476) Point estimates for each bar represent the proportion of patients reporting a given symptom in the deterioration group (n=137) and the no deterioration group (n=339). All participants were included in the calculation of these point estimates. Proportions were calculated by dividing the number of participants reporting a symptom by the total number of patients in each group. Error bars indicate the 95% confidence intervals (95% CI).

**Table 1:** Socio-demographic characteristics of participants reporting symptoms of post-COVID-19 condition (PCC) 3 months after the index emergency department visit, stratified by symptom progression (deterioration versus no deterioration) following vaccination ≥ 90 days post-infection. (N=476)

| Variables<sup>a</sup> | Participants (N=476) |  |  |
| --- | --- | --- | --- |
|  | Deterioration (n=137) | No deterioration (n=339) | P-value |
| Age in years, mean (SD) | 52.4 (15.1) | 54.4 (15.9) | 0.21 |
| Gender<sup>b</sup>, No./total (%) |  |  |  |
| Male | 55/137 (40.1) | 146/339 (43.1) | 0.55 |
| Female | 82/137 (59.8) | 193/339 (56.9) |  |
| Pandemic wave<sup>c</sup>, No./total (%) |  |  |  |
| Pre-Omicron<br>(October 16, 2020 to November 27, 2021) | 136/137 (99.2) | 333/339 (98.2) | 0.39 |
| During Omicron<br>(November 28, 2021 to February 28, 2022) | <5 | 6/339 (1.7) |  |
| Province<sup>c</sup>, No./total (%) |  |  |  |
| British Columbia | 58/137 (42.3) | 108/339 (31.8) | 0.07 |
| Nova Scotia | < 5 | < 5 |  |
| Ontario | 14/137 (10.2) | 23/339 (6.7) |  |
| Québec | 65/137 (47.4) | 206/339 (60.7) |  |
| Saskatchewan | <5 | <5 |  |
| Race<sup>c</sup>, No./total (%) |  |  |  |
| Arab / Middle East | 19/137 (13.8) | 33/339 (9.7) | 0.64 |
| Black | 5/137 (3.6) | 21/339 (6.2) |  |
| East / Southeast Asian | 13/137 (9.4) | 35/339 (10.3) |  |
| Native | <5 | 6/339 (1.7) |  |
| Latin America | 7/137 (5.1) | 13/339 (3.8) |  |
| South Asia | 12/137 (8.7) | 35/339 (10.3) |  |
| White | 69/137 (50.3) | 164/339 (48.5) |  |
| Other | <5 | <5 |  |
| Prefer not to answer | 6/137 (4.3) | 27/339 (7.9) |  |
| Missing data | <5 | <5 |  |
| Level of education<sup>c</sup> , No./total (%) |  |  |  |
| None | 10/137 (7.3) | 14/339 (4.1) | 0.26 |
| High school diploma | 27/137 (19.8) | 82/339 (24.4) |  |
| Vocational certificate or diploma | <5 | 16 |  |
| University certificate or diploma | 12/137 | 25/339 |  |
| University degree or higher | 82/137 (60.2) | 187/339 (55.6) |  |
| Prefer not to answer | <5 | 12/339 (3.5) |  |
| Missing data | <5 | <5 |  |
| Mode of arrival at emergency department, No./total (%) |  |  |  |
| Walk-in | 75/137 (54.7) | 204/339 (60.1) | 0.27 |
| Ambulance | 62/137 (45.2) | 135/339 (39.8) |  |
| Comorbidities<sup>c,d</sup>, No./total (%) |  |  |  |
| Hypertension | 34/137 (24.8) | 112/339 (33.0) | 0.07 |
| Diabetes | 22/137 (16.1) | 63/339 (18.5) | 0.51 |
| Coronary artery disease | 9/137 (6.5) | 21/339 (6.1) | 0.87 |
| Heart failure | <5 | 6/339 (1.7) | 0.39 |
| Chronic kidney disease | <5 | 17/339 (5.0) | 0.16 |
| Mental health diagnosis | 7/137 (5.1) | 15/339 (4.4) | 0.74 |
| Rheumatoid disease | <5 | 25/339 (7.3) | 0.06 |
| Asthma | 16/137 (11.6) | 41/339 (12.1) | 0.89 |
| Chronic liver disease | <5 | <5 | 0.36 |
| Obesity | <5 | 20/339 (5.9) | 0.08 |
| Acute symptoms of COVID-19 during emergency department visit<sup>d</sup>, No./total (%) |  |  |  |
| Cough | 88/137 (64.2) | 210/339 (61.9) | 0.64 |
| Shortness of breath | 88/137 (64.2) | 225/339 (66.3) | 0.65 |
| Fever | 69/137 (50.3) | 171/339 (50.4) | 0.98 |
| Chills | 23/137 (16.7) | 43/339 (12.6) | 0.24 |
| General fatigue | 56/137 (40.8) | 139/339 (41.0) | 0.97 |
| Chest pain | 51/137 (37.2) | 116/339 (34.2) | 0.5 |
| Abdominal pain | 12/137 (8.7) | 29/339 (8.5) | 0.94 |
| Diarrhea | 35/137 (25.5) | 75/339 (22.1) | 0.42 |
| Nausea/vomiting | 30/137 (21.9) | 71/339 (20.9) | 0.81 |
| Headache | 38/137 | 75/339 (22.1) | 0.19 |
| Myalgia | 37/137 (27.01) | 70/339 (20.6) | 0.13 |
| Sore throat | 9/137 (6.5) | 39/339 (11.5) | 0.10 |
| Confusion | 8/137 (5.8) | 17/339 (5.01) | 0.71 |
| Dysgeusia/anosmia | 9/137 (6.5) | 29/339 (8.5) | 0.46 |
| Expectorations | 17/137 (12.4) | 27/339 (7.9) | 0.12 |
| Emergency disposition<sup>c</sup>, No./total (%) |  |  |  |
| Discharge | 91/137 (66.4) | 197/339 (58.1) | 0.24 |
| Admitted | 44/137 (32.1) | 136/339 (40.1) |  |
| Other | <5 | 6/339 (1.7) |  |
| Symptoms of post-COVID-19 condition at the time of vaccination<sup>d</sup>, No./total (%) |  |  |  |
| Post-exertional malaise | 76/137 (55.4) | 197/339 (58.1) | 0.5 |
| Dyspnea | 71/137 (51.8) | 156/339 (46.0) | 0.25 |
| Memory problems | 60/137 (43.8) | 120/339 | 0.08 |
| Concentration problems | 53/137 (38.6) | 98/339 (28.9) | 0.03 |
| Persistent pain | 45/137 (32.8) | 96/339 (28.3) | 0.32 |
| Sleeping difficulties | 43/137 (31.3) | 98/339 (28.9) | 0.59 |
| Other<sup>e</sup> | 36/137 | 82/339 (24.1) | 0.63 |
| Vertigo | 34/137 (24.8) | 68/339 (20.1) | 0.25 |
| Cough | 30/137 (21.9) | 47/339 (13.8) | 0.03 |
| Dysgeusia | 24/137 (17.5) | 43/339 (12.6) | 0.16 |
| Anosmia | 23/137 (16.7) | 55/339 (16.2) | 0.88 |

SD: Standard deviation, No.: Number.
<sup>a</sup> Data on age, sex at birth, province of residence, acute symptoms of COVID-19, mode of arrival and emergency disposition, and pandemic wave were collected from participants' medical records. All other data were collected during the telephone interview with participants.
<sup>b</sup> After analysis, there was no difference between sex and gender in our study.
<sup>c</sup> In accordance with small size restriction policy, numbers less than 5 have not been reported.
<sup>d</sup> Values in this category are not mutually exclusive.
<sup>e</sup> Other symptoms reported consistent with PCC were persistent fatigue, hair loss, anxiety, vision problems.

No significant differences were observed between the two patient groups based on their vaccination status at the time of the index emergency department visit or the number of days between the last pre-infection vaccine dose and the emergency department visit. Receiving a vaccine dose between the index emergency department visit and the 3-month mark confirming a PCC diagnosis was not associated with a difference between groups (Table 2). A higher proportion of participants who received the Moderna vaccine ≥90 days after infection reported a deterioration of their PCC symptoms (38.9%, 95% CI: 30.6-47.3%) compared to those who received the combined AstraZeneca and Pfizer vaccines (25.5%, 95% CI: 20.8-30.2%) (Appendix 4).

**Table 2:** Vaccination status of participants at various periods before receiving the reference vaccine dose ≥ 90 days post-infection and brand of the reference vaccine dose, stratified by symptom progression (deterioration versus no deterioration).

| Variables | Participants, N = 476 |  |  |
| --- | --- | --- | --- |
|  | Deterioration (n=137) | No deterioration (n=339) | P-value |
| Number of SARS-CoV-2 vaccine doses received prior to the emergency department visit, No./total (%)<sup>a,b</sup> |  |  |  |
| 0 | 131 (95.6) | 313 (92.3) | 0.28 |
| 1 | 6 (4.3) | 17 (5.01) |  |
| 2 | <5 | 6 (1.7) |  |
| 3 | <5 | <5 |  |
| Doses received between index emergency department visit and 90 days, No./total (%) |  |  |  |
| 0 | 67 (48.9) | 175 (51.6) | 0.8 |
| 1 | 65 (47.4) | 153 (45.1) |  |
| 2 | <5 | 11 (3.2) |  |
| Brand of vaccine in reference dose ≥ 90 post-infection, No./total (%) |  |  |  |
| AstraZeneca | 5 (3,7) | 12 (3.5) | 0.009 |
| Moderna | 51 (38.1) | 80 (23.4) |  |
| Pfizer | 78 (58.2) | 240 (70.8) |  |
| Missing data | <5 | 7 |  |

SARS-CoV-2: Severe acute respiratory syndrome coronavirus 2, No.: Number.
<sup>a</sup> All vaccine data are self-reported by participants.
<sup>b</sup> In accordance with small size restriction policy, numbers less than 5 have not been reported.

Based on the bivariate analysis, which revealed a significant association between the Moderna (mRNA-1273) vaccine and symptom deterioration (Table 2), the Moderna (mRNA-1273) vaccine was isolated as the primary exposure variable. Due to our small number of available degrees of freedom for the multivariable analysis, the remaining vaccine brands were pooled into a single comparator category, designated as the reference group. This grouping allowed us to effectively test the association of the Moderna vaccine against all other administered vaccine brands, while ensuring sufficient statistical power for the final model.

Our multivariable modeling identified only two independent factors associated with the deterioration of PCC symptoms after a vaccine dose administered ≥90 days after SARS-CoV-2 infection. Receiving a Moderna (mRNA-1273) vaccination dose was associated with an adjusted odds ratio (aOR) of 1.81 (95% CI: 1.15-2.84) (Figure 3) for deterioration compared to other vaccines. A new persistent PCC cough was also associated with the deterioration of PCC symptoms with an adjusted odds ratio of 1.87 (95% CI: 1.06-3.26). No other PCC symptom was associated with PCC symptom deterioration after vaccination. The Hosmer-Lemeshow test yielded a p-value greater than 0.05, suggesting no significant discrepancy between the observed and predicted outcomes and confirming the adequacy of the model adjustment.

**Figure 3.**
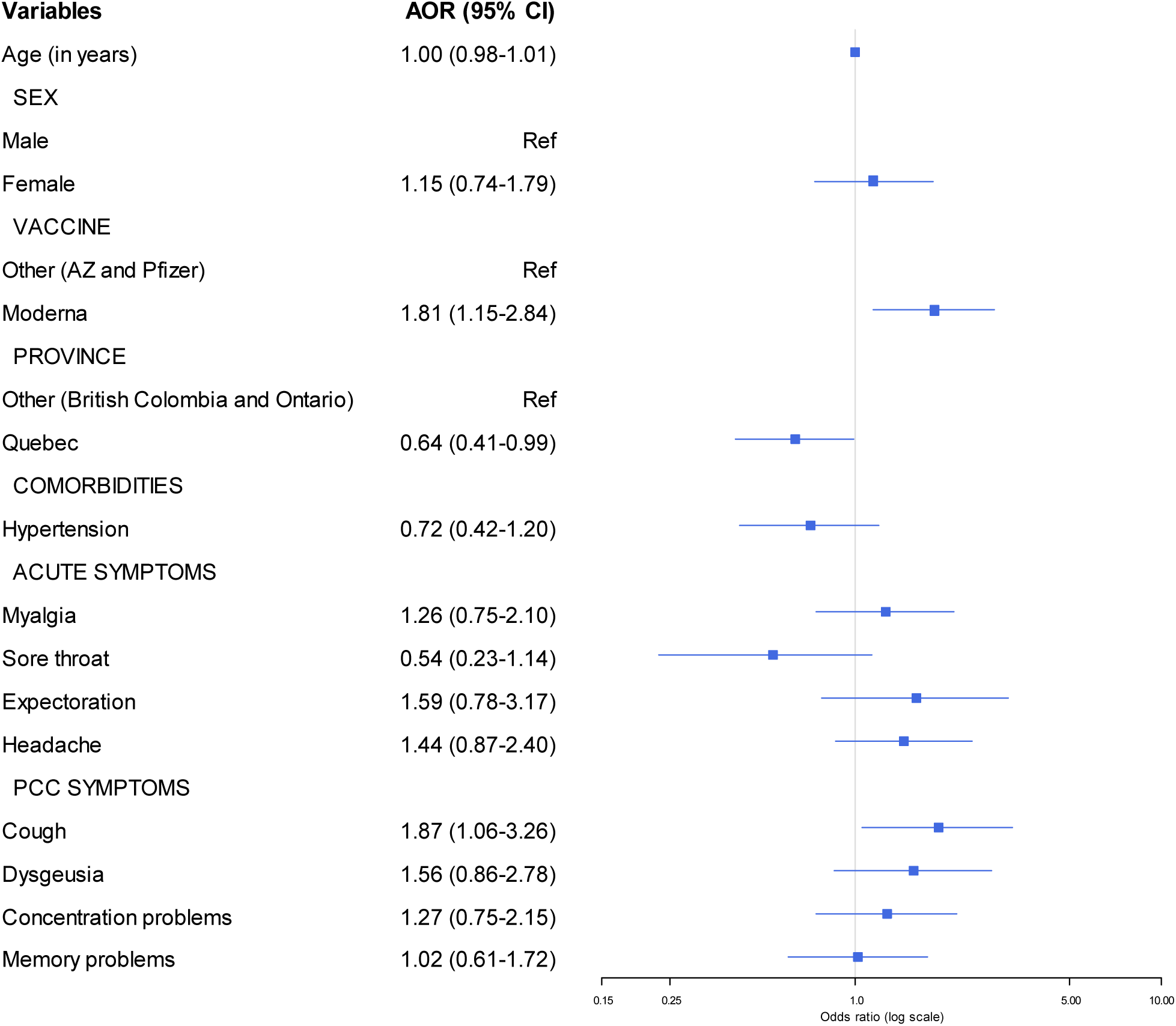
Adjusted odds ratios (aORs) of factors associated with deterioration of post-COVID-19 condition (PCC) symptoms following a SARS-CoV-2 vaccine dose ≥ 90 days after infection (N=476).

We did not find any associations between the sociodemographic characteristics and the deterioration of PCC symptoms. In particular, age and gender were not identified as risk factors.

## Discussion

The objective of our study was to investigate the association between SARS-CoV-2 vaccination and the deterioration of PCC symptoms in patients vaccinated at least 90 days after their initial infection. We found that over a quarter of participants reported a deterioration of their PCC symptoms after receiving a post-infection vaccine dose, while the majority reported no deterioration. In our adjusted model, only two variables were associated with deteriorating symptoms: experiencing a new persistent cough at the time of vaccination and receiving the Moderna (mRNA-1273) vaccine.

The rate of deterioration observed in our study is slightly higher than in comparable studies^20,36^ and much higher than in other studies concluding that symptoms improved or remained stable.^37^ This difference could be attributed to the fact that patients who use emergency departments are in poorer health than the general population ^38^ and have more chronic diseases.^39,40^ Given that PCC is characterized by complex multisystem manifestations with sequelae that can affect almost every organ system, it is reasonable to expect that responses to vaccination may vary and lead to a wide range of outcomes. Similar to other studies on the impact of a vaccine dose on PCC symptoms, sociodemographic characteristics such as age, sex at birth, and race were not associated with change.^36^ Although the symptoms reported by our participants were consistent with those commonly identified in the literature ^41^, the only symptom associated with deterioration in our study was a new persistent cough present at the time of vaccination.

Persistent cough is usually interpreted as a clinical symptom of a chronic inflammatory process in the airways. This type of cough, when it persists for several weeks after a viral infection, is often associated with bronchial hyperreactivity, residual inflammation of the respiratory mucosa, or vagus nerve damage, all of which are mechanisms described in post-viral syndromes, including PCC.^42,43^

In the context of PCC, a persistent cough could indicate that the immune system is still activated in the lungs. Giving an mRNA vaccine, which strongly activates the immune system, could increase this inflammation and worsen other symptoms. The link between an inflammatory symptom at the time of vaccination and later deterioration supports the idea that PCC may involve an immune process that is still active in some patients. ^44,45^ However, it should be noted that we did not identify a link between the presence of asthma and the risk of deterioration, indicating that a history of chronic respiratory disease was not an isolated risk factor for post-vaccination deterioration in patients with PCC.

Another mechanism that could explain the persistence of cough in the context of PCC is related to dysregulation of the renin-angiotensin system (RAS) in the airways. The angiotensin-converting enzyme II (ACE2), widely expressed in the epithelial cells of the respiratory tract, is essential for maintaining this balance by degrading the conversion of angiotensin II, a pro-inflammatory molecule, into angiotensin I, which has anti-inflammatory and vasodilatory properties.^46,47^ During SARS-CoV-2 infection, the virus exploits ACE2 as a receptor to enter the cell, causing its reduction at the cell surface through internalization. The decrease in ACE2 activity leads to an increase in angiotensin II, which exacerbates inflammation in the respiratory tract ^48–51^. This inflammation is known to cause sensitization of sensory nerves in the airways, particularly C fibers, which are involved in triggering the cough reflex. Therefore, the presence of persistent cough could be attributed to both direct viral infection and RAS dysfunction leading to prolonged neuroinflammatory hypersensitivity.

The deterioration of PCC symptoms was associated with a dose of the Moderna (mRNA-1273) vaccine in our study, unlike many other studies that found no difference between vaccines.^18,52^ The vast majority of our participants were vaccinated during the pre-Omicron period and thus received the first version of the vaccines developed against the original strain of COVID-19. Like Pfizer’s (BNT162b2) vaccine, Moderna’s vaccine is an mRNA vaccine encapsulated in lipid nanoparticles that encodes the complete spike protein of the virus.^53^ Since one adult dose of the Moderna (mRNA-1273) vaccine contains three times the number of mRNA copies than the Pfizer vaccine, it has been associated with a greater immune response.^54^ The Moderna vaccine has also been linked to a higher incidence of inflammatory episodes compared to the Pfizer vaccine in certain autoimmune diseases.^55^ In line with the hypothesis that a dysregulated immune response and disruption of anti-inflammatory mechanisms are associated with deteriorating symptoms, this greater humoral and cellular response could explain the difference highlighted in our study.

Some authors have suggested that mRNA vaccines do not exacerbate ACE2 receptor disturbances in individuals with PCC.^49^ Post-vaccination symptoms are attributed to a natural inflammatory response, rather than direct alteration of ACE2 receptors.^49^ According to these authors, the effect of vaccines on these receptors is weak and does not last long, much less than that caused by viral infection. However, this interpretation is controversial. Researchers also believe that the production of the spike protein in respiratory tract cells could reduce the ACE2 receptor. This could cause angiotensin II to become overactive, which could explain symptoms such as persistent coughing. This hypothesis seems to be corroborated by the results of our study, which shows that persistent coughing at the time of vaccination is linked to deterioration symptoms after vaccination. This indicates that there may be more complicated interactions between vaccines and the renin-angiotensin system, and that these should be studied more closely.^56^

Other studies have also found that immune and autoimmune dysfunctions, combined with inflammation, may be exacerbated by vaccination and worsen symptoms such as cough.^57,58^

In addition, there is persistent dysregulation of complement system activation in PCC patients, notably induced by microthrombi formed during the acute phase of COVID-19.^59,60^ In a study that also reported a deterioration in symptoms in nearly a quarter of participants, the deteriorated group had a significantly higher pre-post vaccination antibody titer ratio than the non-deteriorated group, also suggesting an excessive immune response to vaccination is associated with deteriorating symptoms.^61^

It is also interesting to note that although concentration problems were not an explanatory variable in our adjusted model, participants reporting this symptom were significantly more numerous in the deterioration group. This deterioration could be explained by similar physiological processes involving neuroinflammation and immune dysfunction.

Our study has several strengths. First, it is a large, multicenter cohort study conducted in several Canadian provinces among multiple patients with confirmed SARS-CoV-2 infection. By collecting patient-reported data from participants in different provinces, with research assistants who received standardized training, the study identified disparities in certain Canadian geographic areas that warrant further investigation. Second, our study rigorously applied the WHO definition of PCC, specifying precise time thresholds and asking participants to distinguish between new symptoms and chronic symptoms, which enhanced the specificity of participants identified as having PCC and allowed for valid comparisons with other studies. Third, our logistic regression model adjusted for confounding factors allowed us to isolate significant factors associated with symptom deterioration.

Our study has several limitations. First, the effect sizes of the identified variables in our explanatory model are modest with wide confidence intervals due to our small sample size. In addition, we may have missed other explanatory variables for deterioration that could have improved the goodness of fit of our model (e.g., markers of inflammation). However, the model was able to identify clinical variables such as persistent cough to monitor to prevent a potential deterioration of PCC symptoms following a vaccine dose. Second, our questionnaire was implemented without formal psychometric evaluation because there was an urgent need to collect data on PCC at the beginning of the pandemic. However, it was co-developed with patient partners and experts in rehabilitation and PCC, then piloted with a subgroup of patients in French and English and implemented with training to standardize its use. Third, our analyses did not determine whether the reference vaccine brand was homologous or heterologous to the participant’s previous vaccine dose. On average, 54.5% of our participants received at least one dose of vaccine before the reference dose received ≥90 days after infection. If this dose was also Moderna, it is possible that for some participants, a double dose of Moderna was the factor that contributed to the deterioration of symptoms. Fourth, it should be noted that since PCC is a clinical diagnosis based on the exclusion of all other causes, it remains difficult to determine with certainty that the symptoms reported by patients were indeed a new symptom rather than an exacerbation of an underlying condition prior to infection. Another limitation of our study is that we were unable to determine the duration of the deterioration of symptoms after vaccination. We cannot rule out the possibility that the symptoms deteriorated only temporarily, giving way to a more marked improvement. Another limitation of our study was that vaccine data were derived from information provided by the patients themselves. Although patient-reported data have previously been shown to be reliable for vaccine type and timing ^27^, information bias may still exist for this variable.

The results of our study have clinical and public health implications and contribute to knowledge about the impact of vaccination on symptoms of PCC. Although the benefits of COVID-19 vaccination are now indisputable in preventing acute disease and its complications, including PCC, vaccination after a proven episode of infection may be associated with deterioration of persistent PCC symptoms in certain patient profiles. Further research is needed to better understand the mechanisms underlying this variability in vaccination responses and the duration of the deterioration noted in our study. Such research could influence public health recommendations, promote the adoption of an individualized therapeutic approach for certain patient profiles, and potentially lead to the development of tailored vaccine formulations.

## Conclusion

This multi-center study in Canada found that approximately 30% of patients with PCC who received a vaccine dose at least 90 days after infection reported deteriorating symptoms. Patients who received a Moderna (mRNA-1273) vaccine or reported a persistent cough at the time of vaccination were more likely to experience a deterioration of PCC symptoms. These results highlight the need for further understanding of the immuno-inflammatory mechanisms underlying the symptomatic variations observed following vaccination. They also call for an individualized approach to post-infection vaccination strategies, particularly for patients with specific persistent symptoms such as cough.

## Data Availability

Data is available on reasonable request. For investigators who wish to access CCEDRRN data, proposals may be submitted to the network for review and approval by the network s peer-review publication committee, the data access and management committee and the executive committee, as per the network s governance. Information regarding submitting proposals and accessing data may be found on the CCEDRRN website.

https://www.ccedrrn.com/

## Ethical considerations

This study was conducted in accordance with ethical standards for research involving human subjects. It was approved by the research ethics committees of each participating center in the Canadien COVID-19 Emergency Department Rapid Response Network (CCEDRRN, Appendix 2). For the retrospective portion of data collection (data from clinical records), no individual consent was required, in accordance with the authorizations issued by the relevant ethics committees, which deemed that the risks to participants were negligible and that confidentiality was adequately protected. For prospective data collection, participants were contacted by telephone, and verbal informed consent was obtained prior to administering the PCCAQ questionnaire. In the province of British Columbia, in accordance with local guidelines approved by their ethics committee, consent was also permitted to be obtained through a proxy when the participant was unable to respond themselves. All data collected were anonymized and stored on secure servers in accordance with institutional standards for confidentiality and privacy. Finally, Patrick Archambault has received funding from AstraZeneca for a clinical trial unrelated to this study. Patrick Archambault also declares receiving funds from Fonds de recherche du Québec Santé (FRQS). All other authors declare no conflicts of interest.

## Funding

This research was funded by the Canadian Institutes of Health Research (CIHR) (#447679, #464947, and #466880). Malika Seydou Beidari has received support from the following organisations: Long COVID Web (CIHR #185352; Long COVID Web Seed Grant Funding #112707), Centre de recherche du Centre intégré de santé et de services sociaux de Chaudière-Appalaches, The Canadian Network of COVID-19 Clinical Trials Networks (CIHR #458236), VITAM – Centre de recherche en santé durable, and Réseau québécois COVID-19 pandémie.

### Appendix 1: STROBE — Checklist of items that should be included in observational study reports

**Appendix 1: STROBE — Checklist of items that should be included in observational study reports.**
|  | Item No. | Recommendation | Page No. |
| --- | --- | --- | --- |
| Title and abstract | 1 | a) Indicate the type of study using a commonly used term in the title or abstract | 1,2 |
|  |  | b) Provide an informative and balanced overview of what has been achieved and the results obtained in the abstract | 1,2 |
| Introduction |  |  |  |
| Context/justification | 2 | Explain the scientific context and rationale for the reported study | 3 |
| Objectives | 3 | State specific objectives, including any pre-specified assumptions. | 3 |
| Method |  |  |  |
| Study design | 4 | Present the key elements of the study design at the beginning of the article. | 3,4 |
| Background | 5 | Describe the setting, locations, and relevant dates, including recruitment, exposure, follow-up, and data collection periods | 4 |
| Participants | 6 | (a) Indicate the eligibility criteria, as well as the sources and methods of participant selection. Describe the follow-up methods. | 4 |
|  |  | (b) For matched studies, indicate the matching criteria and the number of exposed and unexposed participants | n/a |
| Variables | 7 | Clearly define all outcomes, exposures, predictors, potential confounders, and effect modifiers. Provide diagnostic criteria, if applicable. | 5 |
| Data sources/measurements | 8 | For each variable of interest, indicate the sources of data and details of the assessment methods (measurement). Describe the comparability of assessment methods if there is more than one group. | 5 |
| Bias | 9 | Describe the efforts made to address potential sources of bias. | 18 |
| Sample size | 10 | Explain how the study size was determined. | 4 |
| Quantitative variables | 11 | Explain how quantitative variables were handled in the analyses. If applicable, describe the groupings chosen and justify them. | 5,6 |
| Statistical method | 12 | (a) Describe all statistical methods, including those used to control for confounding factors.<br>(b) Describe all methods used to examine subgroups and interactions.<br>(c) Explain how missing data were handled.<br>(d) If applicable, explain how loss to follow-up was addressed.<br>(e) Describe any sensitivity analyses performed. | 5,6 |
| Results |  |  |  |
| Participants | 13 | (a) Indicate the number of individuals at each stage of the study—for example, the number of people potentially eligible, screened for eligibility, confirmed as eligible, included in the study, completed follow-up, and analyzed.<br>(b) Provide reasons for non-participation at each stage.<br>(c) Consider using a flow diagram. | 6,7 |
| **Descriptive data** | 14 | (a) Provide characteristics of study participants (e.g., demographic, clinical, social) as well as information on exposures and potential confounders.<br>(b) Indicate the number of participants with missing data for each variable of interest.<br>(c) Summarize the duration of follow-up (e.g., mean and total duration). | 6 |
| **Outcome data** | 15 | Report the number of outcome events or summary measures over time. | 7-15 |
| **Main results** | 16 | (a) Provide unadjusted estimates and, where appropriate, estimates adjusted for confounding factors and their precision (e.g., 95% confidence interval). Clearly indicate which confounding factors were considered and why they were included.<br>(b) Indicate the category limits when continuous variables have been categorized.<br>(c) If relevant, consider translating relative risk estimates into absolute risk for a meaningful time period. | 7-15 |
| **Other analyses** | 17 | Report other analyses performed—for example, subgroup and interaction analyses, as well as sensitivity analyses. |  |
| **Discussion** |  |  |  |
| **Key results** | 18 | Summarize the key results with reference to the study objectives. | 15 |
| **Limitations** | 19 | Discuss the limitations of the study, taking into account potential sources of bias or inaccuracy.<br>Address both the direction and magnitude of any potential bias | 18 |
| **Interpret** | 20 | Provide a cautious overall interpretation of the results, taking into account the objectives, limitations, multiple analyses, results of similar studies, and other relevant evidence. | 15-18 |
| **generalizability** | 21 | Discuss the generalizability (external validity) of the study results. | 19 |
| **Funding** | 22 | Indicate the source of funding and the role of funders for this study and, if applicable, for the original study on which this article is based | 19 |

\*Provide information separately for exposed and unexposed groups.

### Appendix 2. Sites name and province of location

Appendix 2. Sites name and province of location.
| Site Name | Province | Start Date | End Date |
| --- | --- | --- | --- |
| Abbotsford Regional Hospital | British Columbia | 2020-12-17 | 2021-05-31 |
| Eagle Ridge Hospital | British Columbia | 2020-12-15 | 2021-05-30 |
| Kelowna General Hospital | British Columbia | 2021-01-04 | 2021-05-25 |
| Lions Gate Hospital | British Columbia | 2021-01-21 | 2022-01-02 |
| Mount St Joseph's Hospital | British Columbia | 2021-01-13 | 2022-01-07 |
| Royal Columbian Hospital | British Columbia | 2020-12-15 | 2021-05-31 |
| Royal Inland Hospital | British Columbia | 2021-02-04 | 2021-05-24 |
| Saint Paul's Hospital | British Columbia | 2021-09-09 | 2022-01-09 |
| Surrey Memorial Hospital | British Columbia | 2020-12-08 | 2021-12-31 |
| Vancouver General Hospital | British Columbia | 2020-12-16 | 2021-09-29 |
| Royal University Hospital | Saskatchewan | 2020-10-29 | 2022-01-03 |
| St Paul's Hospital | Saskatchewan | 2021-04-25 | 2022-01-01 |
| Health Science North Hospital | Ontario | 2021-01-13 | 2022-01-18 |
| Hotel Dieu Hospital | Ontario | 2021-04-16 | 2022-01-23 |
| Kingston General Hospital | Ontario | 2021-01-04 | 2022-02-05 |
| London Health Sciences Center | Ontario | 2021-01-22 | 2021-05-31 |
| Sunnybrook Health Sciences Centre | Ontario | 2021-01-01 | 2022-02-23 |
| The Ottawa Hospital - Civic Campus | Ontario | 2021-03-07 | 2021-05-21 |
| The Ottawa Hospital - General Campus | Ontario | 2023-02-07 | 2021-05-21 |
| Centre Hospitalier de l'Université Laval | Quebec | 2020-12-23 | 2021-01-18 |
| Hôpital de l'Enfant-Jésus | Quebec | 2020-11-28 | 2022-01-31 |
| Hôpital du Sacré-Coeur | Quebec | 2020-12-22 | 2022-02-21 |
| Hôpital du Saint-Sacrement | Quebec | 2021-01-01 | 2021-01-13 |
| Hôpital Saint-François d'Assise | Quebec | 2020-12-29 | 2021-01-09 |
| Hôtel-Dieu de Lévis | Quebec | 2020-10-18 | 2022-02-28 |
| Hôtel-Dieu de Québec | Quebec | 2020-12-29 | 2021-01-05 |
| Institut universitaire de cardiologie et de pneumologie de Québec | Quebec | 2020-12-01 | 2020-12-31 |
| Jewish General Hospital | Quebec | 2020-11-29 | 2021-12-31 |
| Royal Victoria Hospital | Quebec | 2020-12-01 | 2021-11-30 |
| Montréal General Hospital | Quebec | 2020-12-05 | 2021-11-22 |
| Cobequid Community Health Centre | Nova Scotia | 2021-01-08 | 2021-01-15 |
| Dartmouth General Hospital | Nova Scotia | 2021-04-28 | 2022-01-23 |
| Halifax Infirmary | Nova Scotia | 2021-01-26 | 2022-01-24 |

### Appendix 3: Post COVID-19 Condition Assessment Questionnaire

Telephone Follow-up (6-month and 12-month)

Version 2021-11-19

Instructions: The following questions will address the persisting symptoms you might still have after your visit to the emergency department and the utilization of healthcare services.

1. **Did you visit the emergency department since you went on YYYY/MM/DD?**

a. No
b. Yes
c. Prefer not to answer If yes,

i. How many times did you go back to the emergency department? *(number)*
ii. For your first revisit to the ED, do you remember how long it was after your initial visit on YYYY/MM/DD?
  1. A couple of days
  2. Within 1 week
  3. Within 2 weeks
  4. Within 1 month
  5. Between 2 and 6 months
  6. More than 6 months
iii. Were you admitted to the hospital during that visit?
  1. Yes
  2. No
  3. Prefer not to answer
2. **For Research Assistant: Has this participant tested COVID-19 POSITIVE at the Index visit or at any subsequent time?**

a. Yes
b. No If no,

i. Since the beginning of the pandemic, do you believe that you had a COVID-19 infection that was not confirmed by a test?
  1. Yes
  2. No
  3. I don’t know
  4. Prefer not to answer
3. **Since the acute health problem that brought you to the emergency department on YYYY/MM/DD, have you been more short of breath?**

a. No, not at all
b. Yes, and it is still present
c. Yes, sometimes and it is still present
d. Yes, but not anymore
e. I don’t know
f. Prefer not to answer If yes, and it is still present / yes, sometimes and it is still present / yes, but not anymore

i. When do/did you experience shortness of breath?
  1. At rest
  2. During low to moderate intensity activity (e.g., walking, cleaning, doing the dishes)
  3. During high intensity activity (e.g., exercising, jogging, biking) If yes, and it is still present / yes, sometimes and it is still present / yes, but not anymore
ii. When did the shortness of breath start? (calendar) If yes, but not anymore
iii. How long did the shortness of breath last?
  1. A couple of days
  2. 1 week
  3. 2 weeks
  4. Between 1 and 2 months
  5. Between 2 and 6 months
  6. More than 6 months
4. **Since the acute health problem that brought you to the emergency department on YYYY/MM/DD, have you had any new and persisting pain?**

a. No, not at all
b. Yes, and it is still present
c. Yes, sometimes and it is still present
d. Yes, but not anymore
e. I don’t know
f. Prefer not to answer If yes, and it is still present / yes, sometimes and it is still present / yes, but not anymore

i. Where is/was the main source of your pain?
  1. Abdominal pain
  2. Chest pain
  3. Back pain
  4. Sore throat
  5. Stomach ache
  6. Headache or Migraine
  7. Joint pain
  8. Pain while breathing
  9. Pain when coughing
  10. Muscle pain
  11. Pain in the legs
  12. Pain in the arms
  13. Pain when swallowing
  14. Generalized pain (non-specified)
  15. Other
ii. When did the pain start? *(calendar)* If yes, but not anymore
iii. How long did the pain last?
  1. A couple of days
  2. 1 week
  3. 2 weeks
  4. Between 1 and 2 months
  5. Between 2 and 6 months
  6. More than 6 months
5. **Since the acute health problem that brought you to the emergency department on YYYY/MM/DD, have you had a new and persistent cough?**

a. No, not at all
b. Yes, and it is still present
c. Yes, sometimes, and it is still present
d. Yes, but not anymore
e. I don’t know
f. Prefer not to answer If yes, and it is still present / yes, sometimes and it is still present / yes, but not anymore

i. When did the cough start? If yes, but not anymore
ii. How long did the cough last?
  1. A couple of days
  2. 1 week
  3. 2 weeks
  4. Between 1 and 2 months
  5. Between 2 and 6 months
  6. More than 6 months
6. **Since the acute health problem that brought you to the emergency department on YYYY/MM/DD, have you had a decreased sense of smell?**

a. No, not at all
b. Yes, and it is still present
c. Yes, sometimes and it is still present
d. Yes, but not anymore
e. I don’t know
f. Prefer not to answer If yes, and it is still present / yes, sometimes and it is still present / yes, but not anymore

i. When did your decreased sense of smell start? *(calendar)* If yes, but not anymore
ii. How long did your decreased sense of smell last?
  1. A couple of days
  2. 1 week
  3. 2 weeks
  4. Between 1 and 2 months
  5. Between 2 and 6 months
  6. More than 6 months
7. **Since the acute health problem that brought you to the emergency department on YYYY/MM/DD, have you had a decreased sense of taste?**

a. No, not at all
b. Yes, and it is still present
c. Yes, sometimes and it is still present
d. Yes, but not anymore
e. I don’t know
f. Prefer not to answer If yes, and it is still present / yes, sometimes and it is still present / yes, but not anymore.

i. When did your decreased sense of taste start? *(calendar)* If yes, but not anymore
ii. How long did your decreased sense of taste last?
  1. A couple of days
  2. 1 week
  3. 2 weeks
  4. Between 1 and 2 months
  5. Between 2 and 6 months
  6. More than 6 months
8. **Since the acute health problem that brought you to the emergency department on YYYY/MM/DD, has your sleep been different?**

a. No, not at all
b. Yes, and it is still present
c. Yes, sometimes and it is still present
d. Yes, but not anymore
e. I don’t know
f. Prefer not to answer If yes, and it is still present / yes, sometimes and it is still present / yes, but not anymore

i. When did your sleep change? *(calendar)* If yes, but not anymore
ii. For how long was your sleep different?
  1. A couple of days
  2. 1 week
  3. 2 weeks
  4. Between 1 and 2 months
  5. Between 2 and 6 months
  6. More than 6 months
9. **Since the acute health problem that brought you to the emergency department on YYYY/MM/DD, have you felt dizzy or lightheaded?**

a. No, not at all
b. Yes, and it is still present
c. Yes, sometimes and it is still present
d. Yes, but not anymore
e. I don’t know
f. Prefer not to answer If yes, and it is still present / yes, sometimes and it is still present / yes, but not anymore

i. When did your dizziness or lightheadedness start? *(calendar)* If yes, but not anymore
ii. How long did your dizziness or lightheadedness last?
  1. A couple of days
  2. 1 week
  3. 2 weeks
  4. Between 1 and 2 months
  5. Between 2 and 6 months
  6. More than 6 months
10. **Since the acute health problem that brought you to the emergency department on YYYY/MM/DD, has it been harder to concentrate?**

a. No, not at all
b. Yes, and it is still present
c. Yes, sometimes and it is still present
d. Yes, but not anymore
e. I don’t know
f. Prefer not to answer If yes, and it is still present / yes, sometimes and it is still present / yes, but not anymore

i. When did it start to get harder to concentrate? *(calendar)* If yes, but not anymore
ii. For how long was it harder to concentrate?
  1. A couple of days
  2. 1 week
  3. 2 weeks
  4. Between 1 and 2 months
  5. Between 2 and 6 months
  6. More than 6 months
11. **Since the acute health problem that brought you to the emergency department on YYYY/MM/DD, has it been harder to remember things?**

a. No, not at all
b. Yes, and it is still present
c. Yes, sometimes and it is still present
d. Yes, but not anymore
e. I don’t know
f. Prefer not to answer If yes, and it is still present / yes, sometimes and it is still present / yes, but not anymore

i. When did it start to get harder to remember things? *(calendar)* If yes, but not anymore
ii. For how long was it harder to remember things?
  1. A couple of days
  2. 1 week
  3. 2 weeks
  4. Between 1 and 2 months
  5. Between 2 and 6 months
  6. More than 6 months
12. **Since the acute health problem that brought you to the emergency department on YYYY/MM/DD, have you felt unusually tired after physical, mental, or emotional exertion?**

a. No, not at all
b. Yes, and it is still present
c. Yes, sometimes and it is still present
d. Yes, but not anymore
e. I don’t know
f. Prefer not to answer If yes, and it is still present / yes, sometimes and it is still present / yes, but not anymore

i. When did this unusual tiredness start? *(calendar)* If yes, but not anymore
ii. How long did this unusual tiredness last?
  1. A couple of days
  2. 1 week
  3. 2 weeks
  4. Between 1 and 2 months
  5. Between 2 and 6 months
  6. More than 6 months
13. **Since the acute health problem that brought you to the emergency department on YYYY/MM/DD, have you had other symptoms than the ones we discussed that you would like to share today?**

a. No
b. Yes
c. Prefer not to answer If yes,

i. Which one affects/affected you the most?
  1. Anxiety
  2. Behaviour change
  3. Can’t move and/or feel one side of body or face
  4. Constipation
  5. Depressed mood
  6. Diarrhea
  7. Dry mouth
  8. Dysmenorrhea
  9. Dry eyes
  10. Fainting/blackouts
  11. Fever
  12. Food sensitivities
  13. Hot flashes
  14. Jerking of limbs
  15. Joint swelling
  16. Loss of appetite
  17. Loss of interest/pleasure
  18. Loss of teeth
  19. Lumpy lesions: (purple/pink/bluish) on toes/COVID toes
  20. Nausea/vomiting
  21. Night sweats
  22. Numbness or tingling
  23. Persistent fatigue
  24. Hearing problems
  25. Problems passing urine
  26. Problems seeing
  27. Problems swallowing
  28. Problems with balance
  29. Problems with gait/falls
  30. Ringing in ears
  31. Seizures
  32. Skin rash
  33. Slowness of movement
  34. Muscle stiffness
  35. Tremors
  36. Weakness in limbs
  37. Weight loss
  38. Erectile dysfunction
  39. Hallucinations If yes,
ii. When did this symptom start? *(calendar)*
iii. How long did this symptom last?
  1. A couple of days
  2. 1 week
  3. 2 weeks
  4. Between 1 and 2 months
  5. Between 2 and 6 months
  6. More than 6 months
  7. Still present
14. **Since the acute health problem that brought you to the emergency department on YYYY/MM/DD, did you get help from any health services?**

a. No
b. Yes
c. Prefer not to answer If yes,

i. Which health services did you use?
  1. Family doctor
  2. Nurse practitioner
  3. Post-COVID Clinic
  4. Specialist
  5. Physiotherapist
  6. Occupational therapist
  7. Home care nurse
  8. Home care worker
  9. Psychologist
  10. Psychiatrist
  11. Social worker
  12. Respiratory therapist
  13. Pharmacist
  14. Massage therapist
  15. Sex therapist
  16. Alternative medicine specialist (acupuncture, healer, osteopath, homeopath, chiropractor)
  17. Other
15. **Since the acute health problem that brought you to the emergency department on YYYY/MM/DD, are there any health services or health professionals that you were unable to access?**

a. No
b. Yes
c. Prefer not to answer

i. Which health services were you unable to access?
  1. Family doctor
  2. Nurse practitioner
  3. Post-COVID Clinic
  4. Specialist
  5. Physiotherapist
  6. Occupational therapist
  7. Home care nurse
  8. Home care worker
  9. Psychologist
  10. Psychiatrist
  11. Social worker
  12. Respiratory therapist
  13. Pharmacist
  14. Massage therapist
  15. Sex therapist
  16. Alternative medicine specialist (acupuncture, healer, osteopath, homeopath, chiropractor)
  17. Other
16. **Did you notice a change in the symptoms we just talked about after**

a. your first COVID shot?

i. Yes, felt worse after my first shot
ii. Yes, felt better after my first shot
iii. No change
iv. I don’t know
v. No, because I was feeling fine before with no symptoms
vi. I am not vaccinated
vii. Prefer not to answer
b. Did you notice a change in the symptoms after your second COVID shot?

i. Yes, felt worse after my second shot
ii. Yes, felt better after my second shot
iii. No change
iv. I don’t know
v. Did not get a second shot
vi. Prefer not to answer
c. Did you notice a change in the symptoms after your third COVID shot?

i. Yes, felt worse after my third shot
ii. Yes, felt better after my third shot
iii. No change
iv. I don’t know
v. Did not get a third shot
vi. Prefer not to answer
17. **Before the acute health problem that brought you to the emergency department on YYYY/MM/DD, which of the following statements best describes the level of fitness you had then?**

a. Fit and well: you exercised occasionally or regularly and had no medical problems.
b. Managing well: you had some medical problems that limited your activities but didn’t need help
c. Frail: you had medical problems that limited your activities, and needed help with daily activities and personal care
d. I don’t remember
18. **Since the acute health problem that brought you to the emergency department on YYYY/MM/DD, did you receive any disability insurance?**

a. Yes
b. No
c. I don’t know
d. Prefer not to answer

### Appendix 4: Table of participant characteristics by vaccine type received and post-vaccination symptom course (deterioration vs. no deterioration)

|  | Participants (N=466) |  |  |  |
| --- | --- | --- | --- | --- |
|  | AstraZeneca (n=17) | Pfizer (n=318) | Moderna (n=131) | AstraZeneca + Pfizer Combined |
| **Deteriorated, n (%), 95% CI** | 5 (29.4), 8.8-50 | 78 (24.5) 19.8-29.2 | 51 (38.9) 30.5-47.3 | 83 (25.5) 20.8-30.2 |
| **Not deteriorated, n (%), 95% CI** | 12 (70.6) 50-71.2 | 240 (75.5) 70.8-80.2 | 80 (61.1%) 52.7-69.5 | 252 (75.9), 71.3-80.5 |

### Appendix 5: Contributors to the Canadian COVID-19 Emergency Department Rapid Response Network

#### 1. Purpose

This supplementary table provides details of the support staff at each of the participating institutions in the Canadian COVID-19 Emergency Department Rapid Response Network. This supplementary document should be attached to each peer-reviewed manuscript after the methods manuscript (M1). The purpose is to ensure research staffs and lead coordinators are appropriately recognized for their contributions to the network.

#### 2. List of Support Staff

**Table 1.** Network coordinating center staff at the University of British Columbia.

|  |  |  |
| --- | --- | --- |
|  | coordinator | training/onboarding of research assistants. |
|  | coordinator | impact assessment, and qualitative analyses |

**Table 2.**
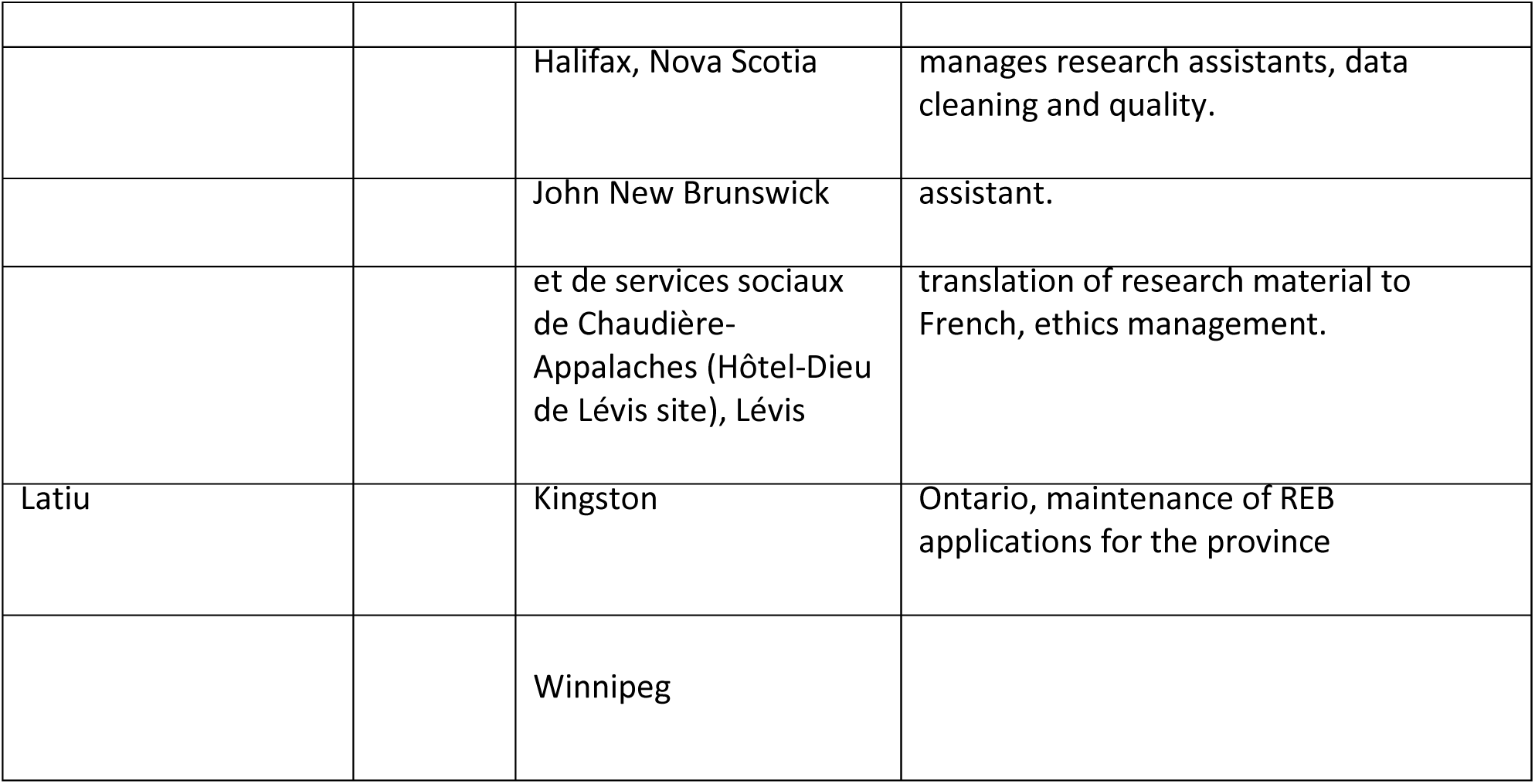
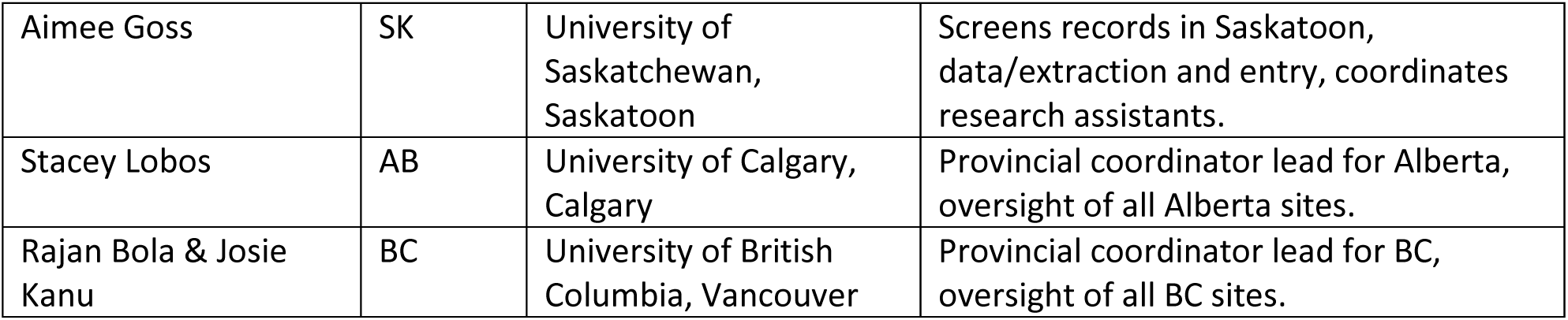
Provincial Coordinators.

**Table 3.** Institutional research assistant (RA) leads. Institutional RA leads are responsible for data extraction and integrity, communication with provincial leads.

| Name | Province | Institutional affiliation(s) |
| --- | --- | --- |
| Corinne DeMone | NS | Dartmouth General Hospital, Cobequid Community Health Centre, Hants Community Hospital<br>Secondary Assessment Centers of the Dartmouth General Hospital, and Halifax Infirmary, Halifax |
| Jacqueline Fraser | NB | Saint John Regional Hospital, Saint John |
| Alexandra Nadeau | QC | CHU de Québec Université Laval, Quebec City |
| Audrey Nolet | QC | Centre intégré de santé et de services sociaux de Chaudière-Appalaches (Hôtel-Dieu de Lévis site), Lévis |
| Xiaoqing Xue | QC | Jewish General Hospital, Montréal |
| David Iannuzzi | QC | McGill University Health Center, Montréal |
| Chantal Lanthier | QC | Hôpital du Sacré-Cœur de Montréal, Montréal |
| Konika Nirmalanathan | ON | University Health Network, Toronto |
| Vlad Latiu | ON | Kingston General Hospital, Hotel Dieu Hospital, Kingston |
| Joanna Yeung | ON | Sunnybrook Health Sciences Center, Toronto |
| Natasha Clayton | ON | Hamilton General Hospital, Juravinski Hospital, Hamilton |
| Tom Chen | ON | London Health Sciences Centre, London |
| Jenna Nichols | ON | Health Sciences North, Sudbury |
| Kate Mackenzie | MB | Health Sciences Centre, Winnipeg |
| Aimee Goss | SK | St. Paul's Hospital, Royal University Hospital, Saskatoon City Hospital, Saskatoon |
| Stacy Ruddell | AB | Foothills Medical Centre, Peter Lougheed Centre, Rockyview General Hospital, South Health Campus, Calgary |
| Natalie Runham | AB | University of Alberta Hospital, Edmonton |
| Karlin Su | AB | Royal Alexandra Hospital/Northeast Community Health Center, Edmonton |
| Taylor Bootsma | BC | St. Paul's Hospital, Mount Saint Joseph, Vancouver |
| Bernice Huynh | BC | Abbotsford Regional Hospital and Cancer Center, Abbotsford |
| Amanda Swirhun | BC | Royal Columbian Hospital, New Westminster |
| Tracy Taylor | BC | Eagle Ridge Hospital and Health Care Centre, Port Moody |
| Mai Hayashi | BC | Royal Inland Hospital, Kamloops |
| Mackenzie Cheyne | BC | Kelowna General Hospital, Kelowna |
| Neenah Williams | BC | Surrey Memorial Hospital, Surrey |
| Katherine Lam | BC | Vancouver General Hospital, Vancouver |
| Kelsey Compagna | BC | Lions Gate Hospital, Vancouver |

**Table 4.**
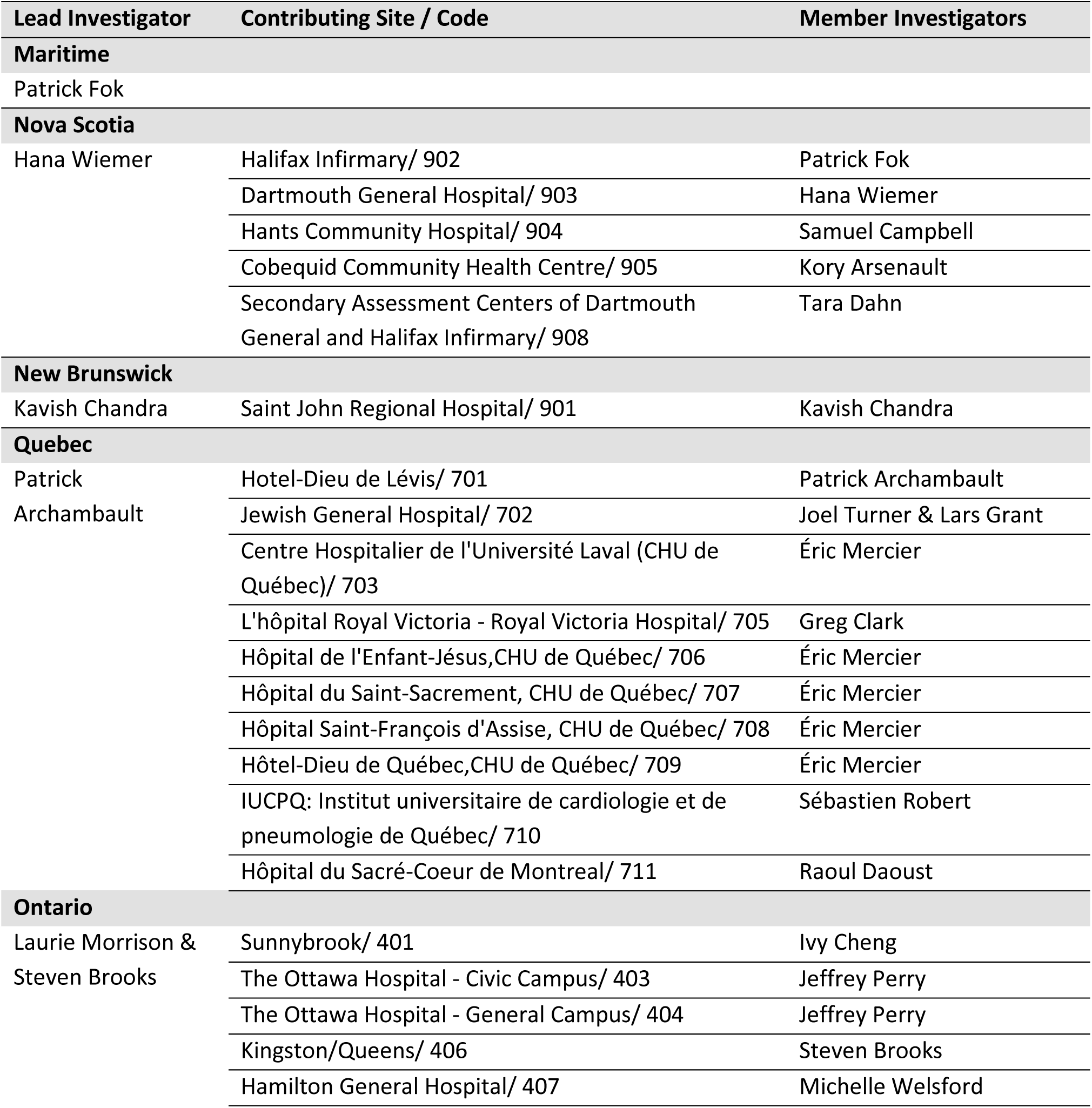
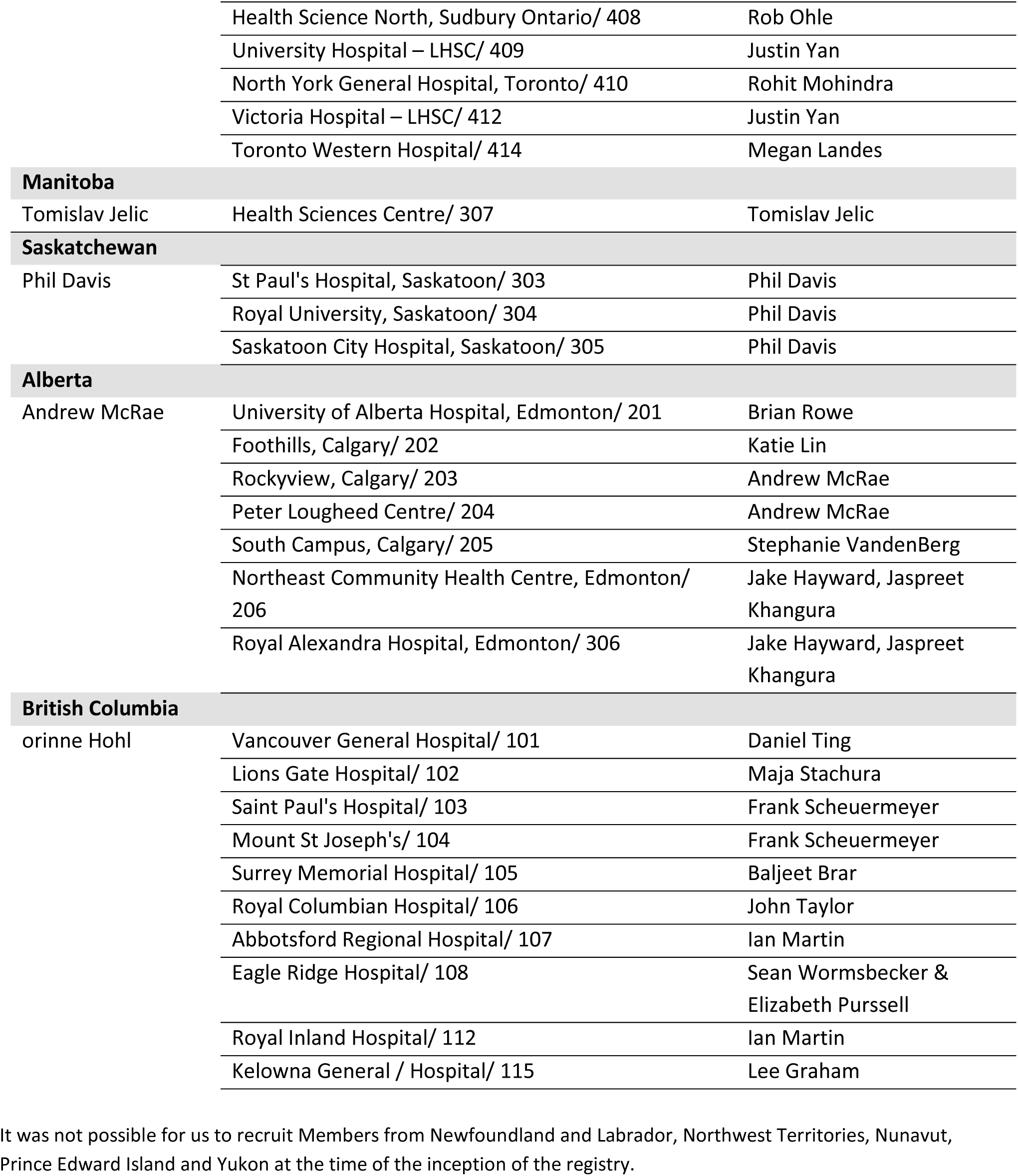
Contributing Study Sites and Investigators.

## Notes

### Funding Statement

This research was funded by the Canadian Institutes of Health Research (#447679, #464947, and #466880). Malika Seydou Beidari has received support from the following organisations: Long COVID Web (CIHR #185352; Long COVID Web Seed Grant Funding #112707), Centre de recherche du Centre intégré de santé et de services sociaux de Chaudiè;re-Appalaches, The Canadian Network of COVID-19 Clinical Trials Networks (CIHR #458236), VITAM Centre de recherche en santé durable, and Réseau québécois COVID-19 pandémie.

### Author Declarations

This study was conducted in accordance with ethical standards for research involving human subjects. This sub-study of the Canadian COVID-19 Emergency Department Rapid Response Network (CCEDRRN), now the Canadian Emergency Department Research Network (CEDRN), was approved by a single, harmonized research ethics board for each participating province. In Canada, for large-scale, multi-site studies, a central ethics review process is utilized. A single ethics committee evaluates the study protocol, and its approval is then recognized by all participating institutions within that jurisdiction. These local institutions only conduct a local review to ensure the protocol aligns with their specific policies and add any necessary site-specific adaptations. This standard practice avoids redundant reviews and streamlines the approval process across multiple research sites. This study was approved by: The University of British Columbia Clinical Research Ethics Board (BC); Nova Scotia Health Research Ethics Board (NS); Comité d’éthique de la recherche du Centre intégré de santé et services sociaux de Chaudiè;re-Appalaches (QC); Queen's University Health Sciences and Affiliated Teaching Hospitals Research Ethics Board (ON); University of Saskatchewan Biomedical Research Ethics Board (SK). For the retrospective portion of data collection (data from clinical records), no individual consent was required, in accordance with the authorizations issued by the relevant ethics committees, which deemed that the risks to participants were negligible and that confidentiality was adequately protected. For prospective data collection, participants were contacted by telephone, and verbal informed consent was obtained prior to administering the PCCAQ questionnaire. In the province of British Columbia, in accordance with local guidelines approved by their ethics committee, consent was also permitted to be obtained through a proxy when the participant was unable to respond themselves. All data collected were anonymized and stored on secure servers in accordance with institutional standards for confidentiality and privacy.

## References

1. World Health Organization. A clinical case definition of post COVID-19 condition by a Delphi consensus, 6 October 2021. https://www.who.int/publications/i/item/WHO-2019-nCoV-Post_COVID-19_condition-Clinical_case_definition-2021.1 (2021).

2. Mudgal, S. K. et al. Long-COVID-19 Impact in non-hospitalized patients: Sleep and quality of life 24 months after SARS-CoV-2 infection. Journal of Family Medicine and Primary Care 13, 1384 (2024).

3. Phetsouphanh, C. et al. Improvement of immune dysregulation in individuals with long COVID at 24-months following SARS-CoV-2 infection. Nature Communications 15, 1–15 (2024).

4. Bowe, B., Xie, Y. & Al-Aly, Z. Postacute sequelae of COVID-19 at 2 years. Nature Medicine 29, 2347–2357 (2023).

5. Thomas, C. et al. Blood Biomarkers of Long COVID: A Systematic Review. Molecular Diagnosis & Therapy 28, 537–574 (2024).

6. Greenhalgh, T., Sivan, M., Perlowski, A. & Nikolich, J. Ž. Long COVID: a clinical update. The Lancet 404, 707–724 (2024).

7. Al-Aly, Z. et al. Long COVID science, research and policy. Nature Medicine 30, 2148–2164 (2024).

8. Gao, P., Liu, J. & Liu, M. Effect of COVID-19 Vaccines on Reducing the Risk of Long COVID in the Real World: A Systematic Review and Meta-Analysis. Int J Environ Res Public Health 19, (2022).

9. Pritchard, E. et al. Impact of vaccination on new SARS-CoV-2 infections in the United Kingdom. Nat Med 27, 1370–1378 (2021).

10. Menni, C. et al. Vaccine side-effects and SARS-CoV-2 infection after vaccination in users of the COVID Symptom Study app in the UK: a prospective observational study. Lancet Infect Dis 21, 939–949 (2021).

11. Harris, R. J. et al. Effect of Vaccination on Household Transmission of SARS-CoV-2 in England. N Engl J Med 385, 759–760 (2021).

12. Lopez Bernal, J., et al. Effectiveness of the Pfizer-BioNTech and Oxford-AstraZeneca vaccines on covid-19 related symptoms, hospital admissions, and mortality in older adults in England: test negative case-control study. BMJ 373, n1088 (2021).

13. Ford, N. D. Long COVID and Significant Activity Limitation Among Adults, by Age — United States, June 1–13, 2022, to June 7–19, 2023. MMWR Morb Mortal Wkly Rep 72, (2023).

14. Kuodi, P. et al. Association between BNT162b2 vaccination and reported incidence of post-COVID-19 symptoms: cross-sectional study 2020-21, Israel. NPJ Vaccines 7, 101 (2022).

15. Wisnivesky, J. P. et al. Association of Vaccination with the Persistence of Post-COVID Symptoms. J. Gen. Intern. Med. 37, 1748–1753 (2022).

16. Peghin, M. et al. Post–COVID-19 syndrome and humoral response association after 1 year in vaccinated and unvaccinated patients. Clinical Microbiology and Infection 28, 1140 (2022).

17. Kim, Y., Bae, S., Chang, H.-H. & Kim, S.-W. Characteristics of long COVID and the impact of COVID-19 vaccination on long COVID 2 years following COVID-19 infection: prospective cohort study. Scientific Reports 14, 1–12 (2024).

18. Ayoubkhani, D. et al. Trajectory of long covid symptoms after covid-19 vaccination: community based cohort study. BMJ 377, e069676 (2022).

19. Babicki, M. et al. Do COVID-19 Vaccinations Affect the Most Common Post-COVID Symptoms? Initial Data from the STOP-COVID Register-12-Month Follow-Up. Viruses 15, (2023).

20. Strain, W. D. et al. The Impact of COVID Vaccination on Symptoms of Long COVID: An International Survey of People with Lived Experience of Long COVID. Vaccines 10, (2022).

21. De Domenico, M. Prevalence of long COVID decreases for increasing COVID-19 vaccine uptake. PLOS global public health 3, (2023).

22. Di Fusco, M. et al. Impact of COVID-19 and effects of booster vaccination with BNT162b2 on six-month long COVID symptoms, quality of life, work productivity and activity impairment during Omicron. Journal of patient-reported outcomes 7, (2023).

23. Hohl, C. M. et al. Development of the Canadian COVID-19 Emergency Department Rapid Response Network population-based registry: a methodology study. Canadian Medical Association Open Access Journal 9, E261–E270 (2021).

24. The Strengthening the Reporting of Observational Studies in Epidemiology (STROBE) Statement: guidelines for reporting observational studies. https://www.equator-network.org/reporting-guidelines/strobe/.

25. Soriano, J. B. et al. A clinical case definition of post-COVID-19 condition by a Delphi consensus. Lancet Infect Dis 22, e102–e107 (2022).

26. World Health Organization. Global COVID-19 Clinical Platform Case Report Form (CRF) for Post COVID condition (Post COVID-19 CRF). https://www.who.int/publications/i/item/global-covid-19-clinical-platform-case-report-form-(crf)-for-post-covid-conditions-(post-covid-19-crf-) (2021).

27. Archambault, P. M., et al. Accuracy of Self-Reported COVID-19 Vaccination Status Compared With a Public Health Vaccination Registry in Québec: Observational Diagnostic Study. JMIR public health and surveillance 9, (2023).

28. Cella, D. et al. Combining Anchor and Distribution-Based Methods to Derive Minimal Clinically Important Differences on the Functional Assessment of Cancer Therapy (FACT) Anemia and Fatigue Scales. Journal of Pain and Symptom Management 24, 547–561 (2002).

29. Cree, B. A. C. et al. Disability improvement as a clinically relevant outcome in clinical trials of relapsing forms of multiple sclerosis. Multiple Sclerosis Journal (2021) doi:10.1177/13524585211000280.

30. Lenz, H.J. et al. Health-related Quality of Life in the Phase III LUME-Colon 1 Study: Comparison and Interpretation of Results From EORTC QLQ-C30 Analyses. Clinical Colorectal Cancer 18, 269–279.e5 (2019).

31. Augustin, M. et al. 15-month post-COVID syndrome in outpatients: Attributes, risk factors, outcomes, and vaccination status - longitudinal, observational, case-control study. Front. Immunol. 14, 1226622 (2023).

32. Domènech-Montoliu, S. et al. Long COVID Prevalence and the Impact of the Third SARS-CoV-2 Vaccine Dose: A Cross-Sectional Analysis from the Third Follow-Up of the Borriana Cohort, Valencia, Spain (2020-2022). Vaccines 11, (2023).

33. Gyöngyösi, M. et al. Effect of monovalent COVID-19 vaccines on viral interference between SARS-CoV-2 and several DNA viruses in patients with long-COVID syndrome. npj Vaccines 8, 1–13 (2023).

34. Hamzaraj, K. et al. Impact of Circulating Anti-Spike Protein Antibody Levels on Multi-Organ Long COVID Symptoms. Vaccines 12, (2024).

35. Archambault, P. M. et al. Post-COVID-19 condition symptoms among emergency department patients tested for SARS-CoV-2 infection. Nature Communications 15, 1–13 (2024).

36. Quach, T. C. et al. Post-COVID-19 Vaccination and Long COVID: Insights from Patient-Reported Data. Vaccines 12, 1427 (2024).

37. Nehme, M. et al. Symptoms After COVID-19 Vaccination in Patients with Post-Acute Sequelae of SARS-CoV-2. J Gen Intern Med 37, 1585–1588 (2022).

38. Giannouchos, T. V., Kum, H.-C., Foster, M. J. & Ohsfeldt, R. L. Characteristics and predictors of adult frequent emergency department users in the United States: A systematic literature review. Journal of Evaluation in Clinical Practice 25, 420–433 (2019).

39. Rosychuk, R. J. et al. Transitions in health care settings for frequent and infrequent users of emergency departments: a population-based retrospective cohort study. BMC Health Services Research 23, 1–12 (2023).

40. Moe, J. et al. People who make frequent emergency department visits based on persistence of frequent use in Ontario and Alberta: a retrospective cohort study. Canadian Medical Association Open Access Journal 10, E220–E231 (2022).

41. Gutzeit, J. et al. Definitions and symptoms of the post-COVID syndrome: an updated systematic umbrella review. European Archives of Psychiatry and Clinical Neuroscience 275, 129–140 (2024).

42. Song, W.-J. et al. Confronting COVID-19-associated cough and the post-COVID syndrome: role of viral neurotropism, neuroinflammation, and neuroimmune responses. The Lancet. Respiratory Medicine 9, 533 (2021).

43. García-Vicente, P., et al. Chronic cough in post-COVID syndrome: Laryngeal electromyography findings in vagus nerve neuropathy. PLoS One 18, e0283758 (2023).

44. Trougakos, I. P. et al. Adverse effects of COVID-19 mRNA vaccines: the spike hypothesis. Trends Mol Med 28, 542–554 (2022).

45. Rocco, J. M. et al. Hyperinflammatory Syndromes After Severe Acute Respiratory Syndrome Coronavirus 2 (SARS-CoV-2) Messenger RNA vaccination in Individuals With Underlying Immune Dysregulation. Clin Infect Dis 75, e912–e915 (2022).

46. Riordan, J. F. Angiotensin-I-converting enzyme and its relatives. Genome Biology 4, 225 (2003).

47. Burrell, L. M., Johnston, C. I., Tikellis, C. & Cooper, M. E. ACE2, a new regulator of the renin-angiotensin system. Trends Endocrinol Metab 15, 166–169 (2004).

48. Patel, S. K. et al. Plasma ACE2 activity is persistently elevated following SARS-CoV-2 infection: implications for COVID-19 pathogenesis and consequences. Eur Respir J 57, (2021).

49. Angeli, F. et al. COVID-19, vaccines and deficiency of ACE2 and other angiotensinases. Closing the loop on the ‘Spike effect’. European Journal of Internal Medicine 103, 23 (2022).

50. El-Arif, G. et al. The Renin-Angiotensin System: A Key Role in SARS-CoV-2-Induced COVID-19. Molecules 26, (2021).

51. Lu, J. & Sun, P. D. High affinity binding of SARS-CoV-2 spike protein enhances ACE2 carboxypeptidase activity. J Biol Chem 295, 18579–18588 (2020).

52. Öztürk, F., Emiroğlu, C. & Aypak, C. The Relationship Between Long Covid Symptoms and Vaccination Status in COVID-19 Survivors. Journal of Evaluation in Clinical Practice 31, e70004 (2025).

53. Baden, L. R. et al. Efficacy and Safety of the mRNA-1273 SARS-CoV-2 Vaccine. The New England Journal of Medicine NEJMoa2035389 (2020).

54. Steensels, D., Pierlet, N., Penders, J., Mesotten, D. & Heylen, L. Comparison of SARS-CoV-2 Antibody Response Following Vaccination With BNT162b2 and mRNA-1273. JAMA 326, 1533–1535 (2021).

55. Fong, W. et al. Prevalence and factors associated with flares following COVID-19 mRNA vaccination in patients with rheumatoid arthritis, psoriatic arthritis and spondyloarthritis: a national cohort study. Advances in Rheumatology 63, 1–9 (2023).

56. Geanes, E. S., McLennan, R., LeMaster, C. & Bradley, T. Autoantibodies to ACE2 and immune molecules are associated with COVID-19 disease severity. Communications Medicine 4, 1–10 (2024).

57. Camici, M., Del Duca, G., Brita, A. C. & Antinori, A. Connecting dots of long COVID-19 pathogenesis: a vagus nerve-hypothalamic-pituitary-adrenal-mitochondrial axis dysfunction. Front. Cell. Infect. Microbiol. 14, 1501949 (2024).

58. Yin, K. et al. Long COVID manifests with T cell dysregulation, inflammation and an uncoordinated adaptive immune response to SARS-CoV-2. Nat Immunol 25, 218–225 (2024).

59. Cervia-Hasler, C. et al. Persistent complement dysregulation with signs of thromboinflammation in active Long Covid. Science (2024) doi:10.1126/science.adg7942.

60. Hawley, H. B. Long COVID: Clinical Findings, Pathology, and Endothelial Molecular Mechanisms. Am J Med 138, 91–97 (2025).

61. Tsuchida, T. et al. Relationship between changes in symptoms and antibody titers after a single vaccination in patients with Long COVID. Journal of Medical Virology 94, 3416–3420 (2022).

